# HYFI: Hybrid filling of the dead-time gap for faster zero echo time imaging

**DOI:** 10.1101/2020.06.16.20132704

**Authors:** Romain Froidevaux, Markus Weiger, Manuela B. Rösler, David O. Brunner, Klaas P. Pruessmann

## Abstract

**Purpose:** To improve the SNR efficiency of zero echo time (ZTE) MRI pulse sequences for faster imaging of short-*T*_2_ components at large dead-time gaps.

**Methods:** HYFI is s strategy of retrieving inner k-space data missed during the dead-time gap arising from radio-frequency excitation and switching in ZTE imaging. It performs hybrid filling of the inner k-space with a small single-point-imaging core surrounded by a stack of shells acquired on radial readouts in an onion-like fashion. The exposition of this concept is followed by translation into guidelines for parameter choice and implementation details. The imaging properties and performance of HYFI are studied in simulations as well as phantom, in-vitro and in-vivo imaging, with an emphasis on comparison with the PETRA technique (pointwise encoding time reduction with radial acquisition).

**Results:** Simulations predict higher SNR efficiency of HYFI compared to PETRA at preserved image quality with the advantage increasing with the size of the k-space gap. These results are confirmed by imaging experiments with gap sizes of 25 to 50 Nyquist dwells, in which scan times for similar SNR could be reduced by 25 to 60%.

**Conclusion:** The HYFI technique provides both high SNR efficiency and image quality, thus outperforming previously known ZTE-based pulse sequences in particular for large k-space gaps. Promising applications include direct imaging of ultra-short *T*_2_ components, such as the myelin bilayer or collagen, *T*_2_ mapping of ultra-fast relaxing signals, and ZTE imaging with reduced chemical shift artifacts.

## 1 Introduction

Direct MRI of tissues with very short transverse relaxation times *T*_*2*_ or *T*_*2*_* in the sub-millisecond range such as e.g. bone (1–3), tendon (4–6), brain (7–10), lung (11–13) and teeth (14–16) is receiving increasing attention due to its potential for both clinical diagnosis and basic research. The rapid signal decay of such tissues prevents detection and spatial encoding through conventional echo-based sequences. Therefore, several dedicated short-T2 techniques have been developed, usually avoiding echo formation (17). One efficient and increasingly used technique is zero echo time (ZTE) imaging (18–21) where a frequency encoding gradient is switched on before radio-frequency (RF) excitation and signal is acquired as soon as possible afterwards (Fig. 1a). In this way, 3D k-space is covered with radial center-out trajectories and spherical support (Fig. 1b).

**Figure 1:**
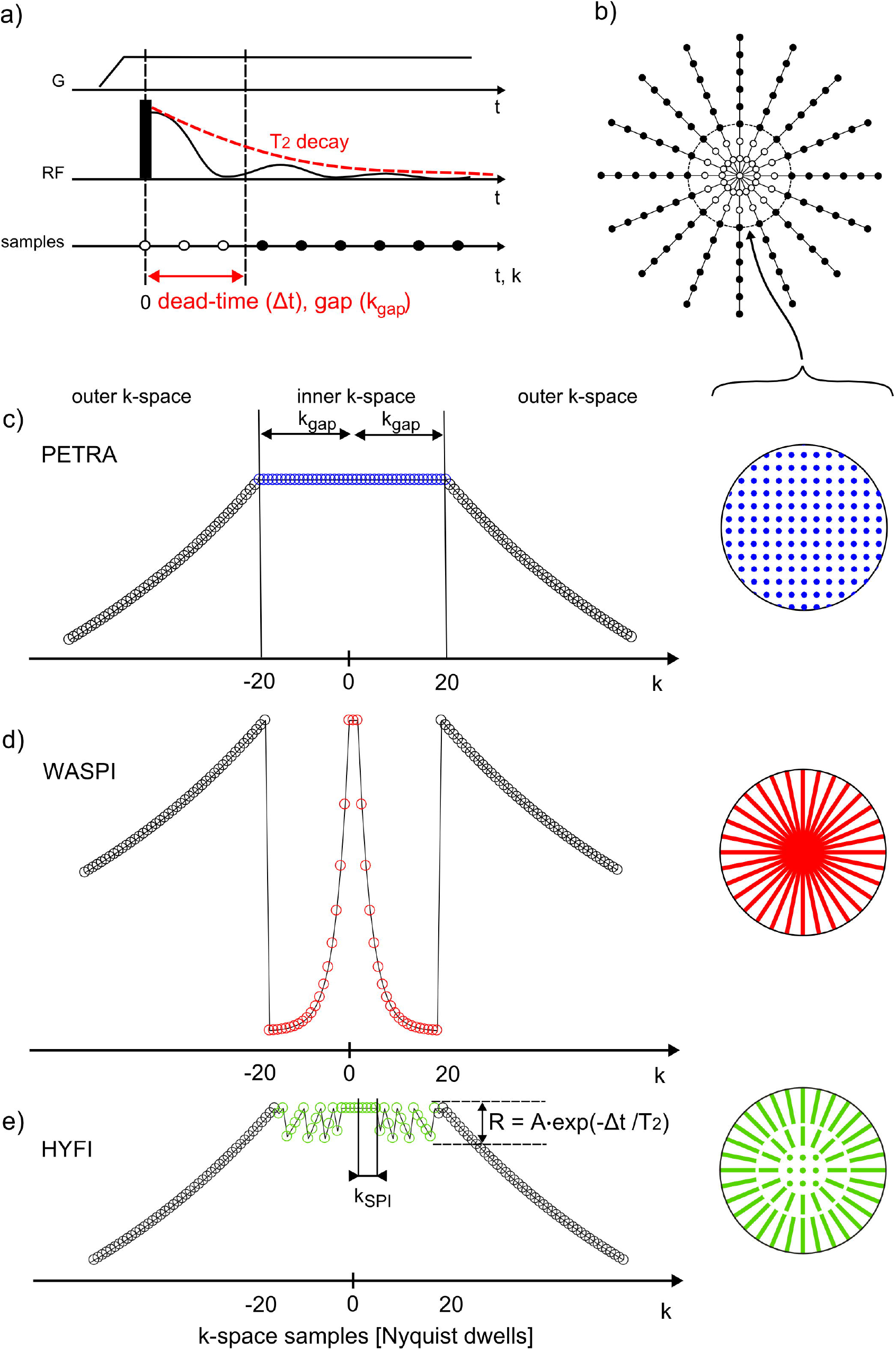
ZTE data acquisition. a) The gradient G applied for radial center-out encoding is ramped up before RF excitation. The initial part of the signal cannot be acquired due to the dead time Δt (white dots). b) After each excitation, a different radial spoke is acquired to fill a 3D k-space volume. The dead time leads to a spherical gap of radius *k*_*gap*_. c-e) PETRA, WASPI, and HYFI provide the data missing in the gap in different ways. Left: 1D depiction of *T*_*2*_ – related MTF in k-space. Right: 2D depiction of inner k-space acquisition geometry. c) In PETRA, the inner k-space is acquired single-point-wise in a Cartesian fashion leading to constant *T*_*2*_ weighting. d) In WASPI, a second set of radial acquisitions is performed at lower gradient strength, giving rise to increased and potentially strong *T*_*2*_ weighting. e) HYFI consists of a Cartesian core (k < *k*_*SPI*_) surrounded by several shells with radial acquisition (k > *k*_*SPI*_). In this way, the *T*_*2*_ decay is restricted to a given range R, wherein R is chosen such as to allow shorter scan time with minimum loss of image quality. Note that the figure partially reuses schemes of previous publications of the same author (26,27).

In ZTE sequences, the dead time Δ*t* caused by RF excitation and switching precludes acquisition of early data, leading to a gap in central k-space (20). No data is available in a sphere of radius *k*_*gap*_ centered on the k-space origin. To avoid related image artifacts, three approaches have been suggested: i) generating the missing information through algebraic reconstruction (22), ii) recovering it through additional acquisitions using Cartesian SPI (single-point imaging) (23,24) as in the PETRA technique (pointwise encoding time reduction with radial acquisition) (21) (Fig. 1c), and iii) recovering the data by radial readouts at lower gradient strength as in the WASPI technique (water- and fat-suppressed proton projection MRI) (19) (Fig. 1d).

Algebraic ZTE has the advantages of not requiring additional acquisition and having a benign behavior of the point spread function (PSF) under T_2_ decay. However, image reconstruction becomes ill-conditioned for *k*_*gap*_ exceeding 3 Nyquist dwells (where a Nyquist dwell is the inverse of the field of view) (25), hence preventing application at larger gaps. WASPI retrieves the missing data with a time-efficient radial acquisition which, however, leads to discontinuous *T*_*2*_-related modulation transfer function (MTF) in k-space and thus a propensity to increased PSF side lobes and associated oscillatory image artefacts (26). On the other hand, PETRA is robust against artifacts but hampered by the slow SPI acquisition of the inner k-space (k < *k*_*gap*_) (26). Therefore, the best sequence choice depends on the particular imaging task and especially on the gap size. However, none of the described methods is well suited for large gaps, i.e., *k*_*gap*_ of tens of dwells, since algebraic ZTE and WASPI lead to poor image quality and PETRA causes undesirably long scan times. Yet, imaging under such conditions can be necessary or beneficial. Indeed, large gaps occur at high imaging bandwidth as required for high-resolution imaging of short-*T*_*2*_ components, in particular in large field of views (27), or when the dead time is relatively large, either because of limitations of the RF hardware or by choice to enable *T*_*2*_ selection (21), *T*_*2*_ mapping, or reduction of chemical-shift artefacts.

In this work, we explore hybrid filling (HYFI) of the k-space center as a path to ZTE imaging that is both time-efficient and robust despite large k-space gaps. The hybrid approach, which reconciles the advantages of point-wise and radial filling, was recently sketched in a conference presentation (28). In the meantime, it has shown promise in a first, challenging applied study (10). On this basis, the present work has the dual aim of establishing the technical basis of HYFI in due detail and assessing its imaging properties and SNR efficiency in comparison with PETRA. This is done by PSF analysis, 3D imaging simulations, and phantom as well as in-vivo imaging.

## 2 Methods

### 2.1 Hybrid filling (HYFI)

The basic idea of HYFI is to replace the SPI part in PETRA by a more time-efficient acquisition strategy with a pattern that still avoids strong discontinuities in the MTF as occurring in WASPI. This is accomplished by hybrid filling of the inner k-space with a small SPI core surrounded by a stack of shells acquired on radial readouts in an onion-like fashion (Fig. 1e). To control image quality, large jumps in signal amplitude associated with *T*_*2*_ decay are avoided by acquiring data over a limited period of time on several radial shells.

Implementation of the described HYFI algorithm is governed by the discrete nature of sampling k-space at Nyquist distance. An example implementation is provided as a Matlab function at the following link. https://doi.org/10.3929/ethz-b-000415045

In more detail, the range R which determines the maximally allowed *T*_*2*_ decay is defined as

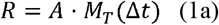

Where

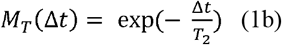

is the amplitude of the transverse magnetization after the dead time, and the decay coefficient

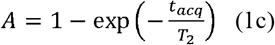

is the proportion of signal amplitude consented to be lost during an acquisition duration *t*_*acq*_. In other words, A describes the size of the range R relative to *M*_*T*_(Δ*t*).

*A* can vary between 0 and 1, corresponding in the limiting cases to PETRA and WASPI, respectively. For a given target *T*_*2*_, *A* is chosen to maximally reduce scan time while keeping artifacts to a negligible level. The latter can be evaluated by means of PSF calculations or image simulations, as shown below. *t*_*acq*_, the maximum acquisition duration allowed to limit the decay of a given *T*_*2*_ to the range R, depends on both the targeted *T*_*2*_ and the value of A. It can be calculated as derived from Eq. (1c) according to

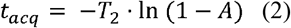

In general, *t*_*acq*_ will be too short to manage acquiring the inner k-space with a single radial spoke. In this case, the inner k-space is split in an onion-like fashion with a core surrounded by one or several shells. Each of these sub-volumes must be acquired within *t*_*acq*_ and starting from Δ*t*. Due to the latter, gradient strengths increase linearly with the distance of the first acquired data point from the origin. Whenever, for a gradient strength, more than one data point can be reached during *t*_*acq*_, radial acquisition is performed as in WASPI to optimize scan time, otherwise SPI is used on a Cartesian grid as in PETRA. As a consequence, the core, where low gradient strengths are required, is probed with SPI while all shells are sampled radially. The core radius is

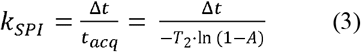

where *k*_*SPI*_ is given in Nyquist dwells. Theoretically *k*_*SPI*_ may reach any value between 0 and _∞_ but is practically rounded to integers and limited to the outer limit of the k-space support (c.f. Matlab function). Moreover, it equals *k*_*gap*_=Δ*t BW*for *A*= *A*_0_, where *A*_0_, the radial onset, is defined by

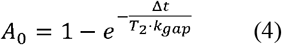

For *A< A*_0_, *k*_*SPI*_ *> k*_*gap*_ and the whole inner k-space is acquired with SPI, thus requiring, 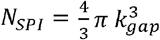number of RF excitations for Cartesian Nyquist sampling. For *A> A*_0_, part of the inner k-space can be acquired radially since *k*_*SPI*_*< k*_*gap*_ and less excitations 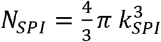 are needed in the core.

As savings in scan time are largely described by the reduction of the SPI core, Fig. 2a illustrates the effects of the choice of *A* on *N*_*SPI*_. In typical cases where Δ*t* < *T*_2_, choosing *A* ≲ 0.1 is sufficient to decrease the size of the SPI core considerably. Moreover, Fig. 2b shows that in such circumstances the PSF lineshape is largely unaffected (More detailed PSFs are shown in the supporting information, Fig. S1), suggesting that scan time can indeed be reduced while preserving image quality.

**Figure 2:**
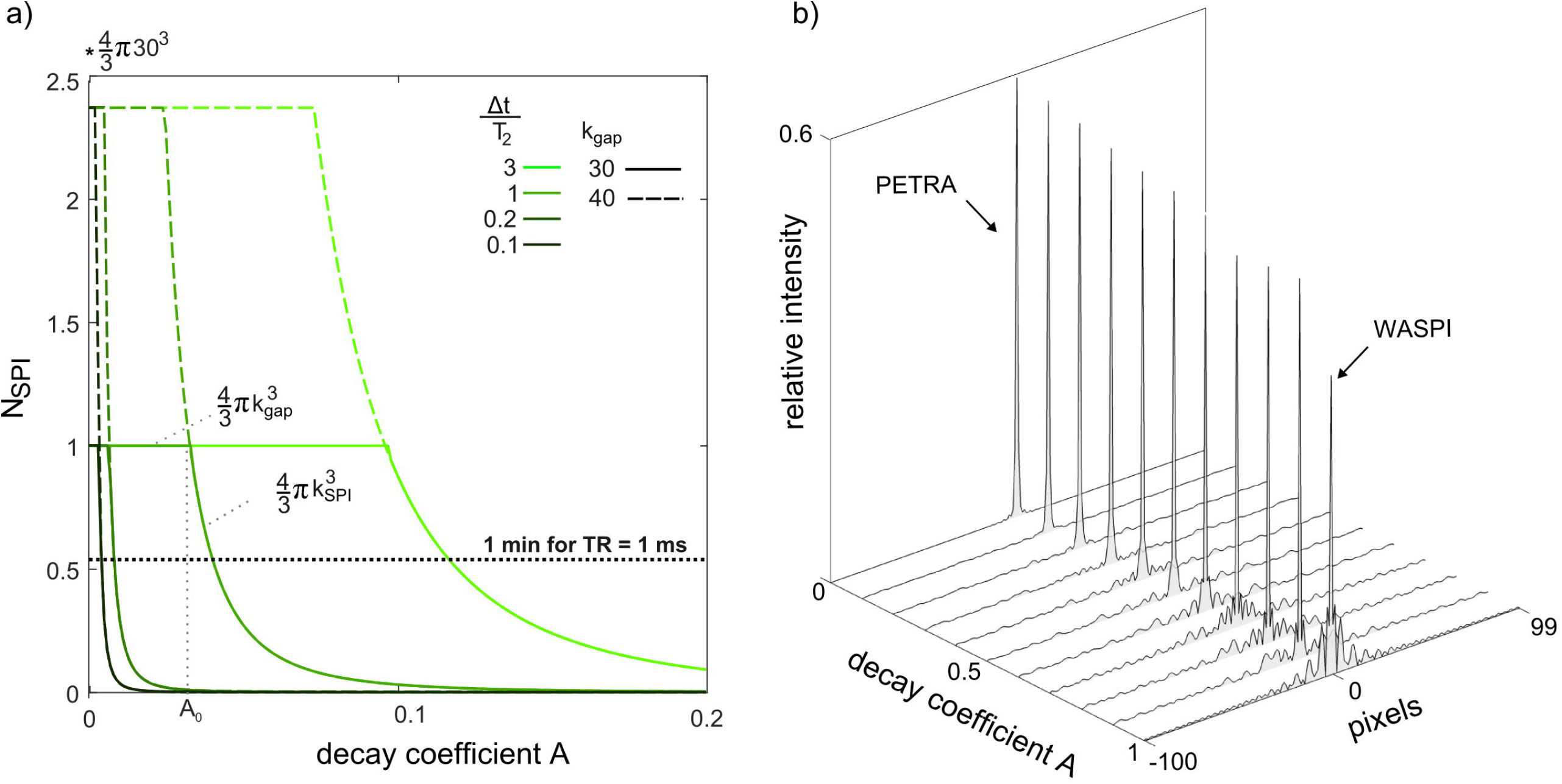
Influence of the decay coefficient *A* on scan time and depiction fidelity. a) Number of excitations required to fill the SPI core (N_SPI_) as a function of the decay coefficient *A* for different ratios Δ*t*/*T*_2_ and gap sizes k_gap_. For given gap and Δ*t*/*T*_2_ ratio, as *A* increases, N_SPI_ stays constant up to a particular value *A* (Equ. 4) and then starts diminishing. In more detail, when *A* is small, the complete inner k-space must be acquired single-point-wise since *k*_*SPI*_ > *k*_*gap*_ and hence 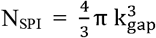. However, for larger *A*, when *k*_*SPI* ≤_ *k*_*gap*_, part of the inner k-space can be acquired radially. Hence the SPI core is limited to the sphere of radius *k*_*SPI*_ and 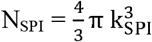. In summary, this figure shows that for common situations where Δ*t* is smaller or comparable to *T*_*2*_, the volume of data that needs to be acquired in a time-consuming SPI manner rapidly decreases to almost negligible values even with relatively small decay coefficient. Assuming a TR of 1 ms this allows reducing scan time by several minutes compared to PETRA. b) Point spread function lineshapes. Calculations were done assuming *T*_*2*_ = 100 Nyquist dwells and *k*_*gap*_ *= 30*. As *A* increases from 0 (PETRA) to 1 (WASPI), the main lobe amplitude decreases and side lobes build up. Importantly, the PSF lineshapes are well preserved for *A*≈ 0.1 at which N_SPI_ is substantially decreased. Hence, HYFI is expected to reduce scan time while preserving image quality. More details PSFs are shown in Fig. S1. Note that PETRA and WASPI acquisitions are also obtained with values of A slightly larger 0 and smaller than 1 respectively (c.f. Matlab script for more details).

Around the core, in each radial shell the angular spoke density is adapted to fulfill the Nyquist criterion at the outer shell radius to minimize scan time. The possible savings in scan time for the complete inner k-space including the acquisition of both core and shells are illustrated in Fig. 3. In the limiting cases of PETRA and WASPI, the number of RF excitations, N_gap_, required to fill a sphere of radius *k*_*gap*_, varies with the volume and the surface of the sphere respectively. In HYFI, N_gap_ depends on the decay coefficient *A* and is bounded by these limits. As already suggested in Fig. 2, small decay coefficients are sufficient to significantly decrease N_gap_.

**Figure 3:**
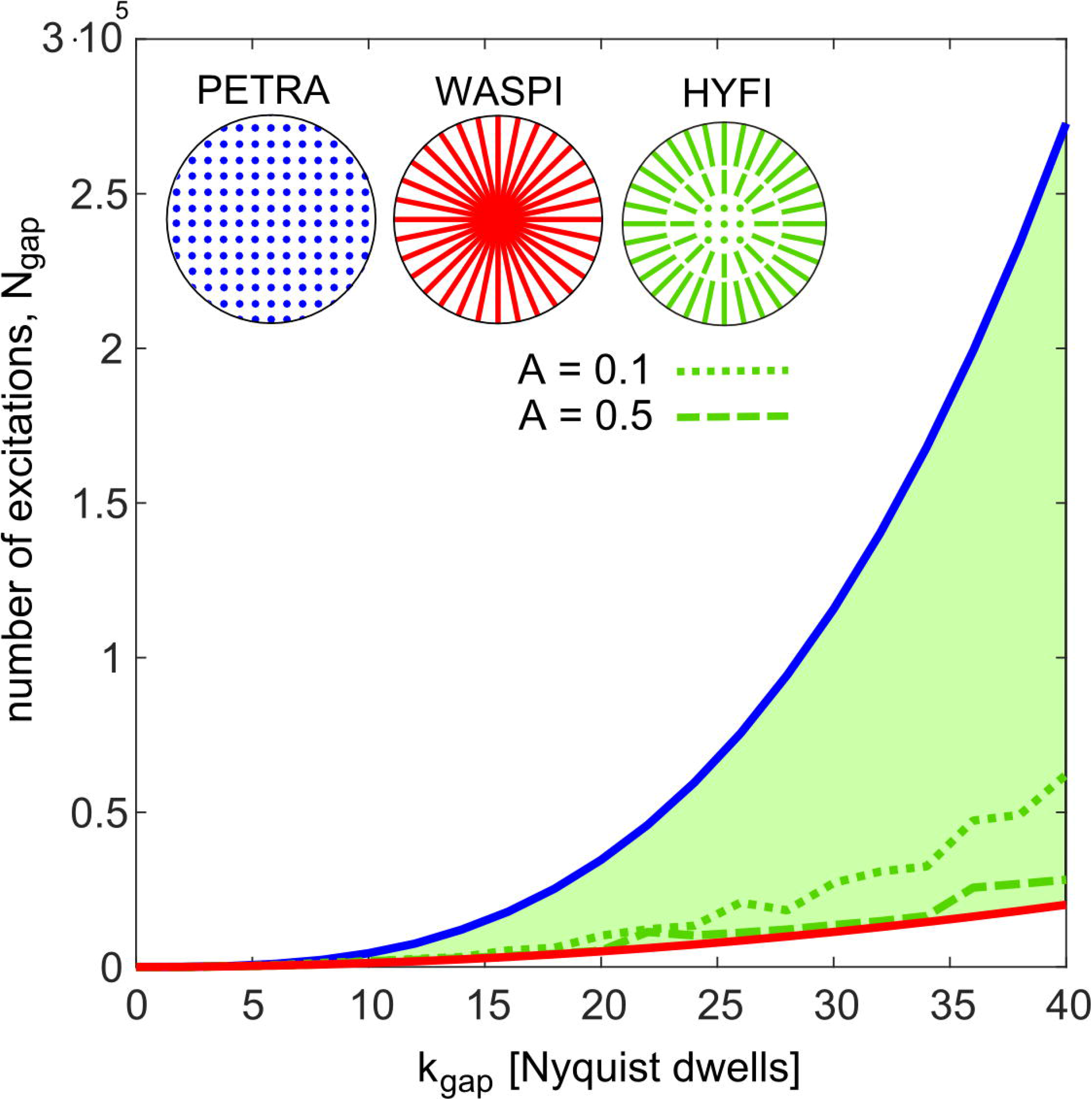
Number of excitations required to fill the complete inner k-space by the different techniques. A decay with *T*_*2*_ = 100 Nyquist dwells is assumed. Circles at the top illustrate the acquisition geometries. The number of excitations, *N*_*gap*_, required to fill the inner 3D k-space evolves with 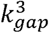 for PETRA and 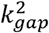 for WASPI. In the proposed HYFI method, inner k-space is filled by a combination of SPI and radial acquisitions, and thus the green area enclosed by the curves for PETRA and WASPI becomes accessible. Green lines represent selected HYFI acquisitions with decay coefficients A = 0.1 and 0.5.

### 2.2 Choice of imaging parameters

All imaging parameters are listed in Tab. 1.

Generally, small dead times are desired to maximize signal amplitude and reduce scan duration. However, in some situations it may be preferred to deliberately extend the dead time and/or the gap size. For example, short-*T*_*2*_ components that cannot be resolved may be selectively suppressed in this way. Moreover, in the inner k-space, data samples are acquired in a small time range bounded by the shell acquisition duration t_acq_. When t_acq_ is small enough, this leads to similar *T*_*2*_-weighting and chemical shift-induced phase for all inner k-space data. As a consequence, increasing the inner k-space volume to a substantial part of the support improves the accuracy of *T*_*2*_ mapping based on a series of such data. In addition, it also reduces PSF blurring (27) and as well as chemical shift artifacts. In this work, increased dead time is employed for suppression of components with extremely short *T*_*2*_, reduction of chemical shift artifacts, as well as *T*_*2*_ mapping.

When setting up a protocol, values for target *T*_2_ and decay coefficient *A* must be selected. A suitable choice of the parameter pair (*A, T*_2_) is crucial for optimal HYFI performance. Figure 2 reveals that a good compromise between image quality and scan time is obtained for *A* ≲ 0.1. However, in most cases the imaged samples contain multiple signal sources with different relaxation times *T*_2_. Then a choice of *A* which is appropriate for a given *T*_2_ leads to stronger decay for signals with shorter *T*_2_ and hence potentially to artifacts. As usually not all relaxation times present in a sample are known a priori, an educated choice of the target *T*_2_ may not always be possible. As a rule of thumb, in the presence of multiple *T*_*2*_s it is considered safe to choose a target *T*_*2*_ = Δ*t* since signals with *T*_*2*_ < Δ*t* will considerably decay before data acquisition. Moreover, as shown in the results section, the target *T*_*2*_ can be chosen larger than Δ*t* if the MR signal is dominated by sources with *T*_*2*_ >> Δ*t*.

### 2.3 Hardware

All experiments were performed on Achieva MRI systems (Philips Healthcare, Best, Netherlands) at 3 T or 7 T complemented with symmetrically biased transmit-receive switches (29) with switching times of approximately 3 µs at 3 T and 1 µs at 7 T, custom-made spectrometers (30) with up to 4 MHz acquisition bandwidth and short digital filters with group delays down to 1.2 µs. Moreover, the 3 T scanner was equipped with a high-performance gradient insert system capable of reaching 200 mT/m at full duty cycle (31) and a broadband linear RF power amplifier BLA1000-I E (Bruker Biospin, Wissembourg, France) with the advantage of a relatively small ring-down signal (32). Largely 1H-free RF coils were used for both transmission and reception, a surface coil of 80 mm diameter and two birdcage coils (33,34). Block and sweep HSN pulses with bandwidth matching the imaging bandwidth were used for RF excitation.

### 2.4 Samples

An imaging phantom with two different *T*_*2*_s was created by placing a stack of erasers with *T*_*2*_ ≈ 300 µs onto a disk made of rubber with *T*_*2*_ ≈ 100 µs.

A bone phantom was taken from a previous study (26). The piece of bovine tibia of 60 mm diameter and 25 mm thickness had been freed of sources of long-lived MR signal, i.e. soft tissues. The signal of the bone is dominated by two *T*_*2*_s of about 10 and 150 µs. Hence, during the imaging process, a relatively long dead time of 40 µs was chosen deliberately to suppress the shorter component that cannot be resolved to the targeted sub-millimeter resolution and to focus on the longer-*T*_*2*_ contributions, as well as to increase the PSF-limited resolution (27).

In-vivo imaging of a knee and a head was conducted in healthy volunteers according to applicable ethics approval, and written informed consent was obtained from all subjects. For knee imaging, the dead time was intentionally increased to 200 *µs* to extend the inner k-space and thus reduce chemical shift artifacts.

An imaging phantom with a range of *T*_*2*_ values was created by filling six solutions of MnCl_2_ with concentrations of 240, 120, 60, 30, 15 and 7.5 mMol into glass vials. For measuring the transverse relaxation times, the solutions were filled into glass spheres of 20 mm diameter to minimize susceptibility effects. Free induction decay (FID) signals were acquired and fitted with single exponentials, providing decay constants of 54, 92, 181, 341, 663 and 1271 µs respectively. For *T*_2_ mapping, a series of images was acquired with constant gap (*k*_*gap*_ = 47) and different dead times (Δ*t* = 55, 100, 200, 400, 600 µs). The signal decay was fitted exponentially with the echo time chosen as *TE=* Δ*t*.

### 2.5 Image reconstruction and processing

Images were reconstructed using an iterative conjugate gradient algorithm (35) complemented by sampling density calculation (36) and pulse correction for modulated pulses (37).

For determining the SNR in images, additional noise data was acquired in the absence of RF excitation. The average signal over a region of interest (ROI) in the magnitude sample image was divided by the standard deviation of the noise image in the same ROI. The SNR efficiency was obtained as 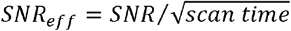. Finally, the relative scan time for equal SNR was calculated according to

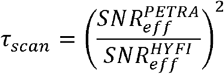

When interpreting a series of images acquired at different dead time Δ*t*, the echo time is defined as TE = Δ*t* for both PETRA and HYFI.

### 2.6 Simulations

The k-space signal of spheres with 50, 30, 18 and 9 mm diameter and *T*_*2*_s of 100 µs was created analytically (38) and images were reconstructed with the algorithm described above.

## 3 Results

The effect of the decay coefficient *A* on image quality and scan efficiency is illustrated with 3D simulations (Fig. 4). Corresponding image profiles are shown in Fig. S2. As the decay coefficient *A* increases, the relative number of excitation needed to fill the inner-k-space, n_gap_, diminishes, increasing the scan efficiency. For this sample, images without noticeable artifacts are obtained with A < 0.1.

**Figure 4:**
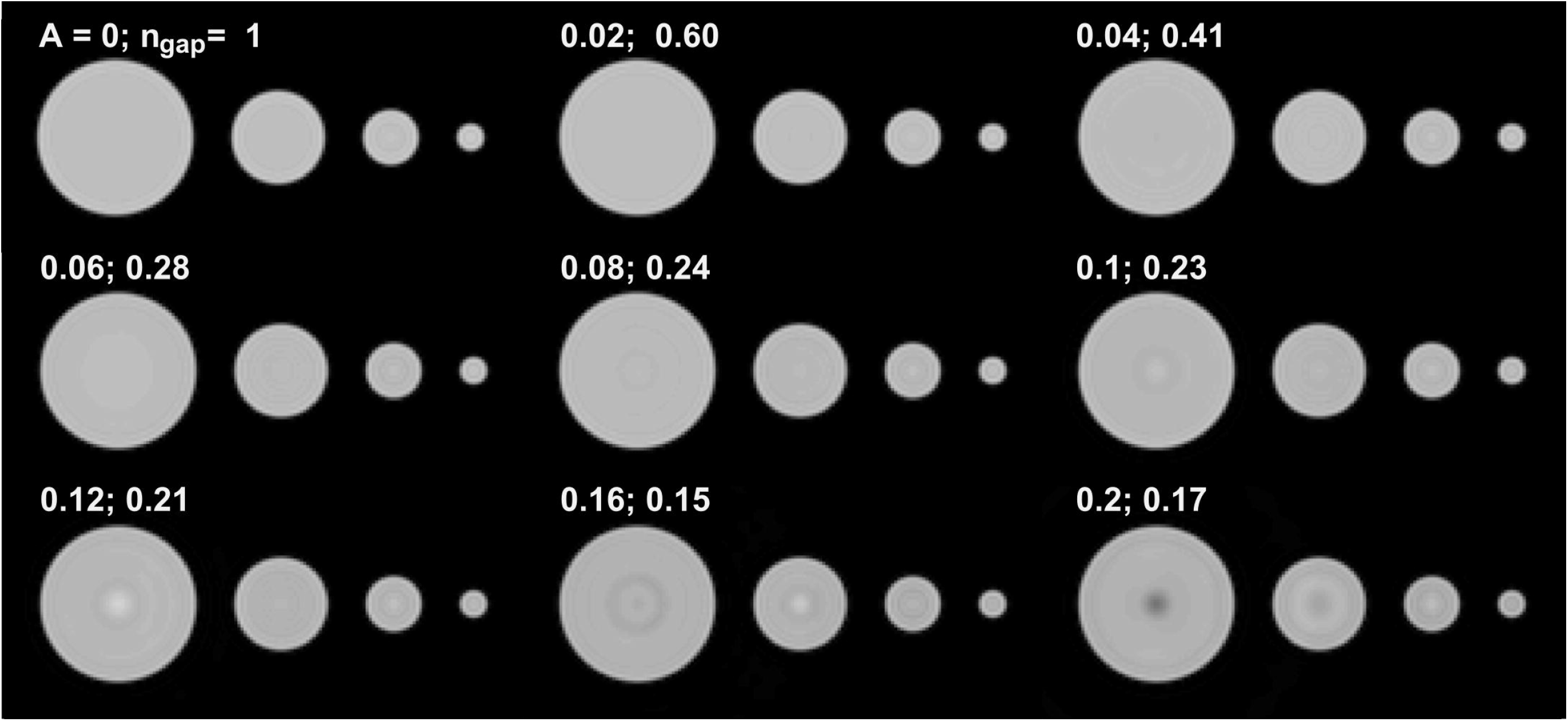
3D HYFI simulations illustrating the effect of the amplitude coefficient A on image quality and scan efficiency. Imaging of spheres with 50, 30, 18 and 9 mm diameter was simulated assuming *T*_*2*_ = 100 µs and *k*_*gap*_ = 30 Nyquist dwells. Other parameters are listed in Table 1. As *A* increases, *k*_*SPI*_ decreases and the relative number of excitations needed to acquire the inner k-space, *n*_*gap*_, decreases. It quickly reaches 28% (*A* = 0.06) with preserved image quality. Above this value, *n*_*gap*_ decreases slower and artifacts start to appear in the center of the larger spheres, suggesting that for such samples the optimal decay coefficient *A* resides below 0.1. Image profiles of a few representative cases are shown in Fig. S2.

The phantom experiment in Fig. 5 demonstrates the HYFI principle over a large range of *A*. For *A* = 0, the inner k-space is acquired in an SPI fashion, leading there to a constant plateau of *T*_2_ weighting, maximum *n*_*gap*_ and artifact-free images. When *A* increases, radial shells with restricted decay replace part of the SPI plateau and *n*_*gap*_ diminishes. However, this also creates progressively increasing irregularities in the MTF which in turn lead to increasingly large ringing artifacts out- and inside the imaged object.

**Figure 5:**
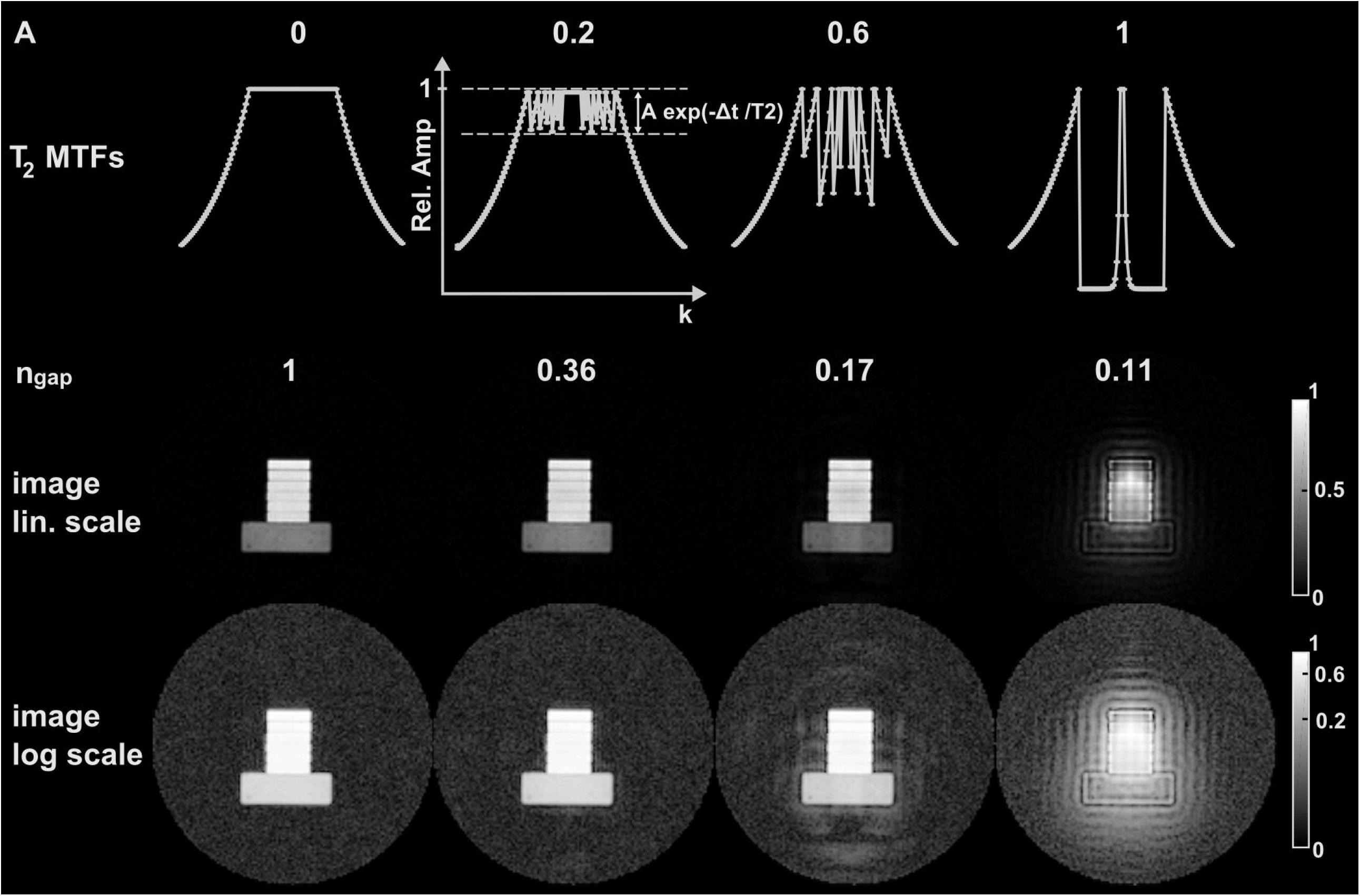
Demonstration of HYFI principle in a phantom experiment. The first row represents the decay coefficient *A* growing from 0 to 1. Below are the corresponding 1D k-space *T*_*2*_ *–*related MTFs and the relative number of excitations required to fill the gap, n_gap_. The resulting images are shown in the two bottom rows with linear and logarithmic greyscales.

**Table 1:** Parameter table. The most important scan parameters are the target *T*_*2*_, the bandwidth BW, the field of view FOV, the gradient G, the voxel size Δ*r*, the matrix size M, the dead time Δ*t*, the k-space gap *k*_*gap*_, the repetition time TR, the number of sample averages NSA, the pulse type, the pulse duration, the coil type and the scanner field strength.

| Sample | $T_2$ | BW<br>[kHz] | FOV<br>[mm] | G<br>[mT/m] | $\Delta r$<br>[mm] | M | $\Delta t$<br>[μs] | $k_{gap}$<br>[dw] | TR<br>[ms] | NSA | Pulse | Pul.<br>dur<br>[μs] | Coil | B0 [T] |
| --- | --- | --- | --- | --- | --- | --- | --- | --- | --- | --- | --- | --- | --- | --- |
| Sphere simulations | 100 | 1000 | 200 | 117.4 | 1 | 200 | 30 | 30 | x | x | x | x | x | x |
| Stack of erasers | 100 | 250 | 240 | 24.5 | 1.9 | 128 | 100.0 | 25 | 1 | 1 | Block | 2 | Birdcage | 7 |
| Bone | 200 | 766 | 90 | 199.9 | 0.4 | 256 | 40.0 | 31 | 1 | 8 | Block | 2 | Surface | 3 |
| Head | 200 | 2000 | 300 | 156.60 | 1.7 | 176 | 15.0 | 30 | 0.65 | 19 | HSN | 8 | Birdcage | 3 |
| Knee | 500 | 250 | 240 | 24.50 | 1.0 | 240 | 5.5 | 2 | 1 | 1 | Block | 2 | Birdcage | 7 |
| Knee | 500 | 250 | 240 | 24.50 | 1.0 | 240 | 200.0 | 50 | 1 | 1 | Block | 2 | Birdcage | 7 |
| MnCl<sub>2</sub> vials | 55 | 851.52 | 100 | 199.90 | 1.0 | 100 | 55.0 | 47 | 1 | 1 | HSN | 10 | Surface | 3 |
| MnCl<sub>2</sub> vials | 100 | 470 | 100 | 110.38 | 1.0 | 100 | 100.0 | 47 | 1 | 1 | HSN | 10 | Surface | 3 |
| MnCl<sub>2</sub> vials | 200 | 235 | 100 | 55.20 | 1.0 | 100 | 200.0 | 47 | 1 | 1 | HSN | 10 | Surface | 3 |
| MnCl<sub>2</sub> vials | 400 | 117.5 | 100 | 27.60 | 1.0 | 100 | 400.0 | 47 | 1 | 1 | HSN | 10 | Surface | 3 |
| MnCl<sub>2</sub> vials | 600 | 78 | 100 | 18.32 | 1.0 | 100 | 600.0 | 47 | 1 | 1 | HSN | 10 | Surface | 3 |

In Fig. 6, the performance of PETRA and HYFI is compared for imaging a sample of bovine tibia. Both techniques lead to high quality images depicting fine trabecular bone structure. Other details appear in the maximum intensity projection, such as glue from the coil conductor and the fixation tape. In HYFI, the choice of a small *A* = 0.04 is sufficient to significantly improve the SNR efficiency compared to PETRA, which translates in a 25% decrease of total scan time for same SNR.

**Figure 6:**
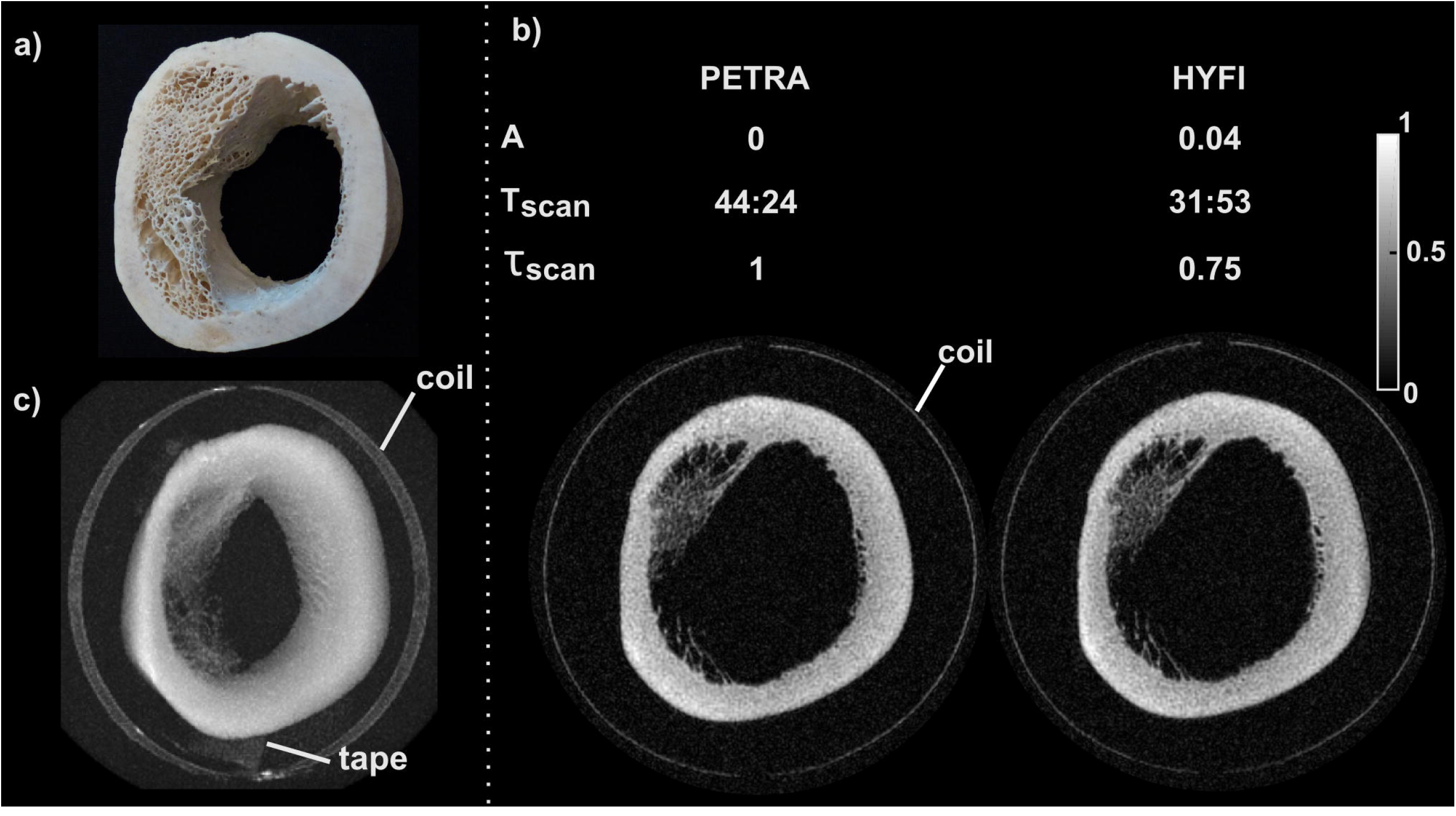
Bone imaging. a) Picture of the imaged segment of a bovine tibia (Here will be a statement saying that this picture is reproduced from ref 27 as soon the permission is received from Wiley). b) Comparison of PETRA and HYFI, with the decay coefficient *A*, the total scan time T_scan_ in min:sec and the relative total scan time for same SNR, *τ*_*scan*_ (including both inner and outer k-space). c) Maximum intensity projection of the HYFI image demonstrating high image quality over the whole field of view. The glue fixing the copper coil to the glass support and the tape holding the sample to the bed are also depicted.

The head images with an unusually high bandwidth of 2 MHz presented in Fig. 7 confirm the above results. The SNR efficiency of HYFI is enhanced compared to PETRA and lead to 40% scan time reduction for same SNR while preserving image quality.

**Figure 7:**
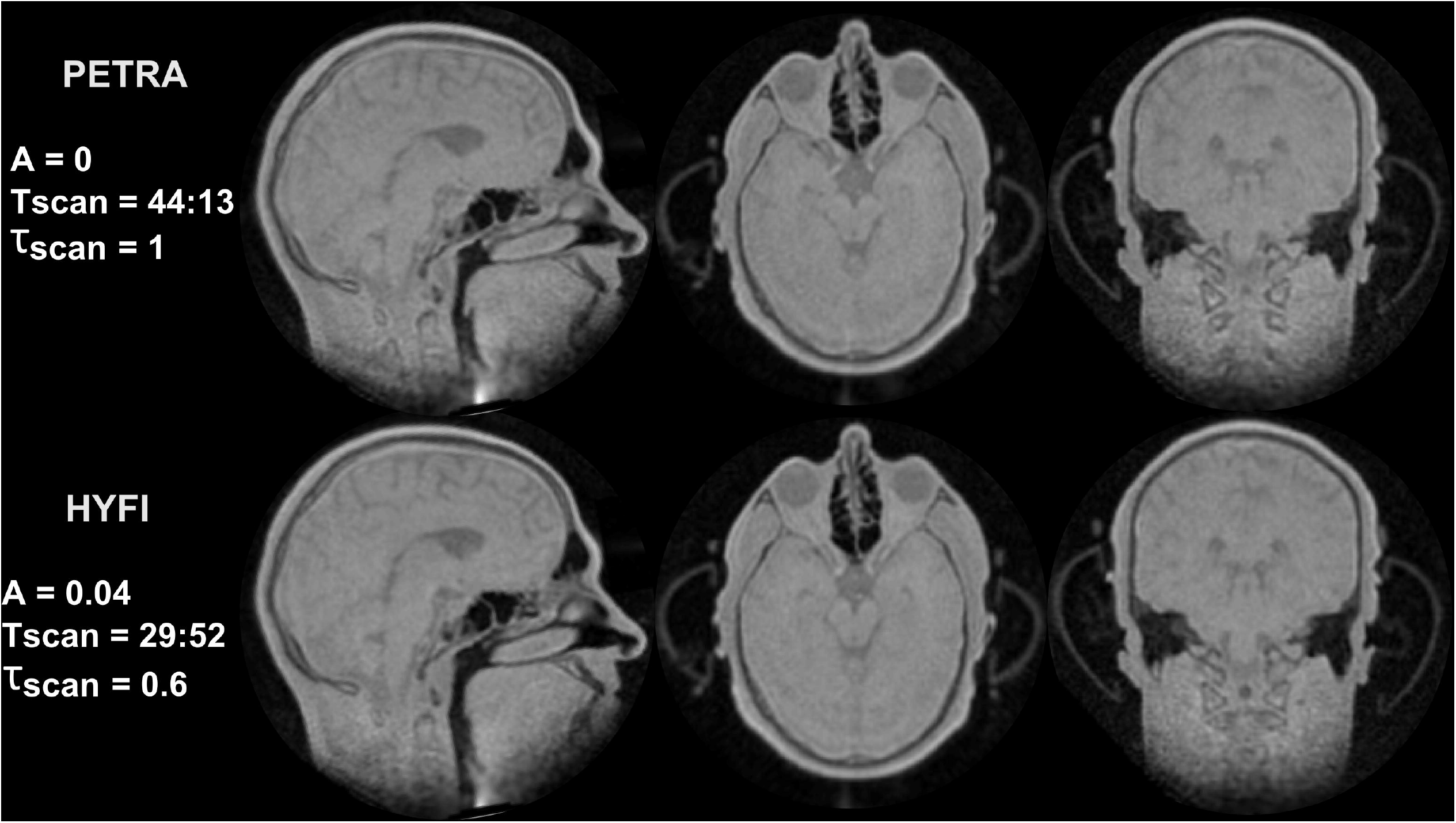
High gradient head imaging. The displayed parameters are the decay coefficient *A*, the total scan time T_scan_ in min:sec and the relative scan time for same SNR, *τ*_*scan*_. Top row: Three perpendicular slices of the same 3D volume acquired with PETRA. Bottom row: The corresponding HYFI images acquired with considerably reduced *τ*_*scan*_.

Figure 8 shows the influence of the dead time gap on chemical shift artifacts in ZTE knee imaging. At minimum dead time Δ*t* = 5.5 µs leading to *k*_*gap*_= 1.4 Nyquist dwells, signal intensity is maximized and moreover, the missing data points can be reconstructed algebraically leading to minimum scan duration. However, due to signal dephasing during the encoding duration of 480 µs, chemical shift artefacts appear at water-fat boundaries (17). Increasing the dead time to 200 µs, enlarges the inner k-space to 50 Nyquist dwells radius and thus reduces the acquisition time range for data located in this region. In PETRA (Fig. 8b), signal can dephase only within 1 Nyquist dwell = 4 µs. Hence, while accepting a loss of signal intensity associated to the longer dead time, chemical shift artifacts are strongly reduced and resolution at water-fat interfaces is improved. However, scan time is substantially increased. In the same circumstances, HYFI provides similar image quality but reduces the acquisition duration for same SNR by 62% as compared to PETRA (Fig. 8c).

**Figure 8:**
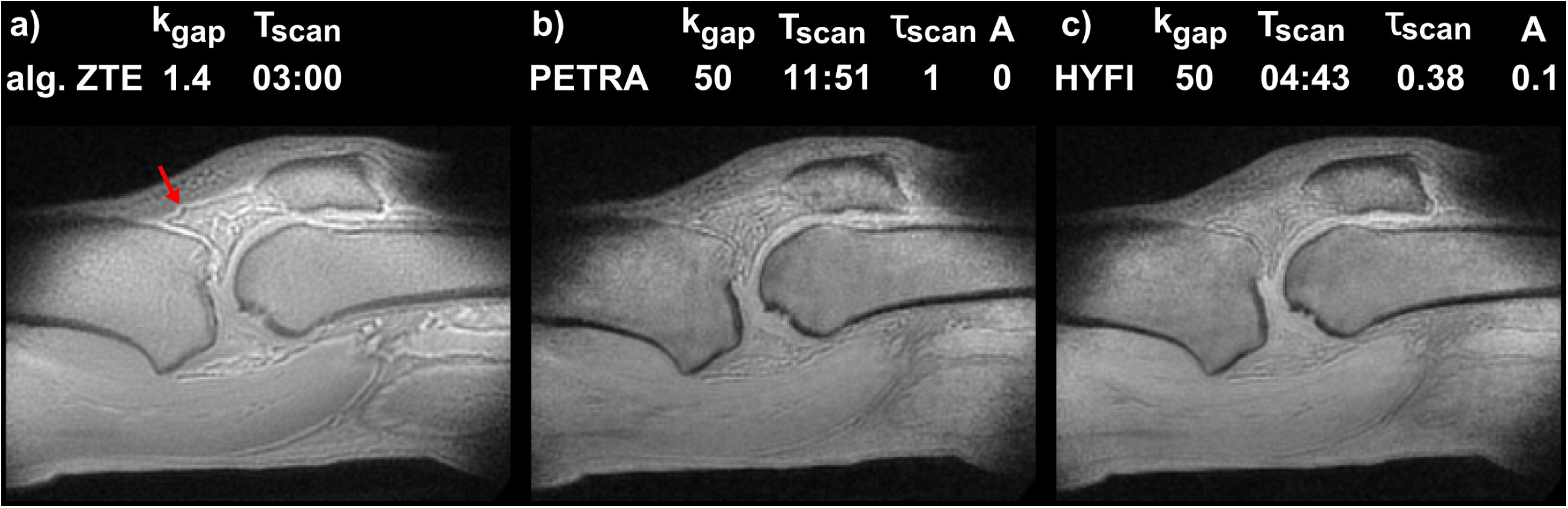
Knee imaging. a) Algebraic ZTE imaging with short dead time and hence small gap, short scan time, but relatively strong chemical shift artefacts at water-fat boundaries as indicated by the red arrow. b) PETRA image with gap intentionally increased to 50 Nyquist dwells to acquire most of the data within a reduced time range and hence decrease chemical-shift artefacts c) Same as in b) except that the inner k-space is filled with HYFI (A = 0.1). In this case the relative scan time for same SNR (*τ*_*scan*_) decreases by 62% while image quality is preserved.

*T*_*2*_ mapping of short-*T*_*2*_ samples is demonstrated in Fig. 9. To enable accurate fitting, the inner k-space was deliberately increased up to the outer limit of the support such that the whole k-space was acquired within a restricted time range. In this way, PETRA approaches pure SPI acquisition (24). Figure 9a shows that at *TE* = 55 µs all vials are well depicted and image intensity drops at larger *TE* in the short-*T*_*2*_ samples. For same image and *T*_*2*_ map quality, the scan time of HYFI is 62% lower than PETRA. Figure 9b shows a good correspondence between the fit of average map intensities and the fit of the FID, especially in the short-*T*_*2*_ range. However, two observations can be made: 1) There is an increasing divergence between FIDs and *T*_*2*_ maps as *T*_*2*_ gets larger, and 2) the relaxation times fitted from the HYFI data are slightly but consistently smaller than the PETRA results. The first observation is assigned to residual B0 inhomogeneity in the samples used for FID measurements, leading to smaller effective *T*_*2*_* values. The second effect results from the definition of the echo time *TE=* Δ*t* which suits PETRA better than HYFI. In the radial shells of HYFI, data is acquired for a duration t_acq_ following Δ*t*. During that time, the signal decays and appears smaller than in PETRA, therefore generating a reduction in image intensity that increases with *TE* and thus leads to smaller fitted *T*_*2*_-values.

**Figure 9:**
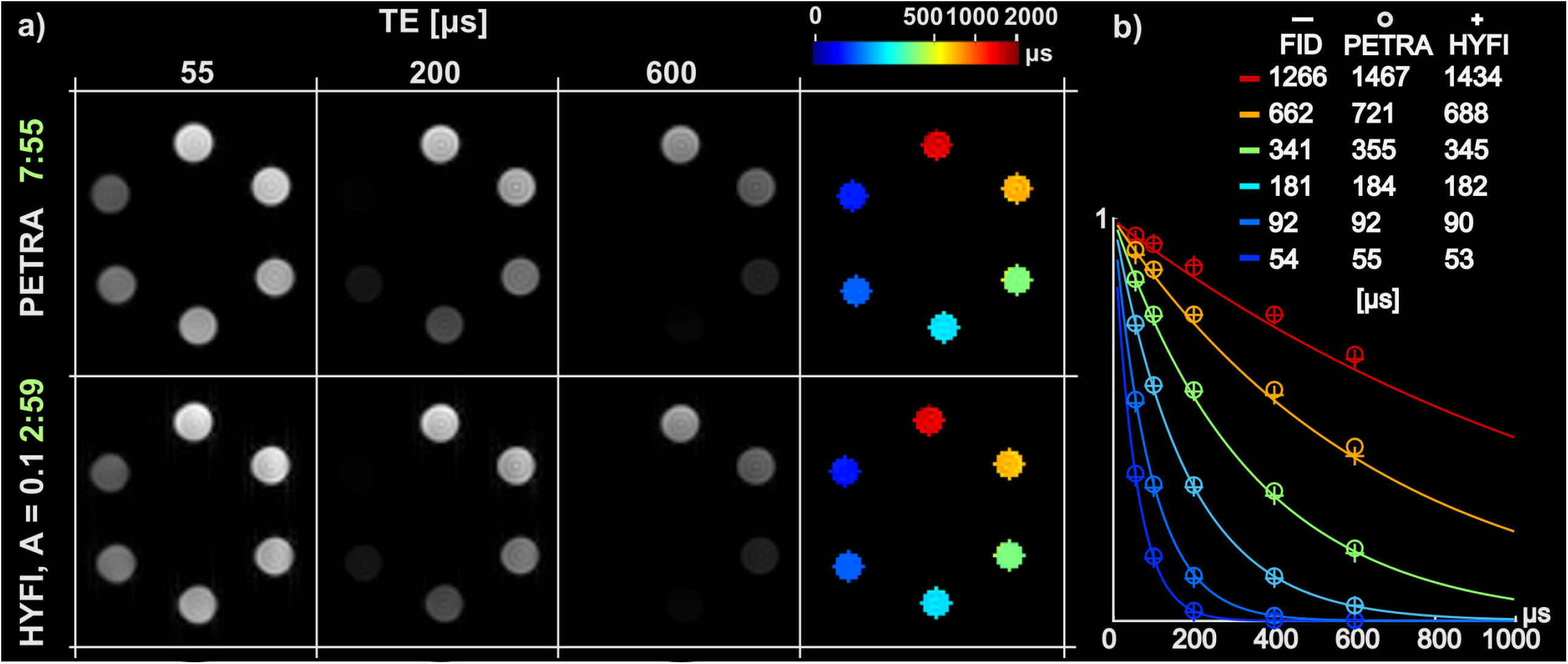
Short-*T*_*2*_ mapping in MnCl_2_ solutions. a) Series of PETRA and HYFI images taken at different echo time *TE=*Δ*t* = 55, 100, 200, 400, 600 µs (only a subset is displayed). The maps of the transverse relaxation times were obtained by pixel-wise fitting the signal decay (last column). The scan time of individual images is given in min:sec (green) which shows a considerable reduction for HYFI. b) The relative intensities of the signal averaged over each sample are plotted as a function of *TE* (circles and crosses) and overlaid with the free induction decay (FID) measured in each MnCl_2_ solution (solid line). Fitted single-exponential *T*_*2*_ values are given in the legend.

## 4 Discussion

HYFI, a recently introduced ZTE-based method with hybrid filling of the inner k-space was described in detail and its performance in the presence of large k-space gaps was studied. It was demonstrated that with HYFI, substantial reductions in scan time can be enabled while preserving image quality as compared to the PETRA technique. The advantage of HYFI increases with gap size and is therefore of particular interest at high bandwidth, large minimum RF dead times, and large dead times selected to manipulate image contrast. The technique was successfully employed for imaging on different phantoms and in vivo.

Using HYFI with high efficiency and fidelity necessitates a suitable choice of the parameter pair (*A, T*_2_). The simulations in Fig. 4 demonstrate that in the presence of a single *T*_*2*_, *A* ≲ 0.1 provides considerable savings in scan time at still high image quality. However as *A* is increased, artifacts become more likely due to coherent interaction of increasingly large PSF side lobes, occurring predominantly in the center of large objects.

In more common cases involving different tissues and molecules and thus a range of transverse relaxation times, the choice of the target *T*_2_ requires extra considerations. Selecting the smallest *T*_2_ present in the sample is safe but usually too conservative. Typically, components with *T*_2_< Δ*t* have little influence on the final image and choosing a target *T*_2_= Δ*t* can be considered an appropriate rule of thumb (Fig. 9). Moreover, the target *T*_2_ might be increased to values larger than Δ*t* without degrading image quality if the signal is dominated by components with longer *T*_2_. In the presented in-vivo data, target *T*_2_s of a few hundreds of microseconds still lead to images without noticeable artifacts (Figs. 7 and 8) although components with clearly faster relaxation but lower intensity, e.g. myelin or collagen, are present. Only if the latter signals should be extracted from the data, they need to be considered for setting the target *T*_2_.

In the performed experiments, the reductions in scan time (for same SNR) of HYFI with respect to PETRA range from 25% to 62% (Figs. 6-8). The influencing factors are gap size, spatial resolution, and the parameter pair (*A, T*_2_). The relative number of excitations required to fill the inner k-space with HYFI decreases with increasing gap size when compared to PETRA (Fig. 2). This explains why the best HYFI performance is obtained in Figs. 8 and 9, were gaps were deliberately increased to high values to reduce chemical shift artefacts and perform *T*_2_ mapping, respectively. The resolution determines the time spent for the acquisition of the outer k-space which in turn affects the relative scan time spent on the inner k-space. Thus, as resolution and hence scan time are increased, the absolute time difference between PETRA and HYFI does not change but the relative advantage of HYFI diminishes. Finally, the selection of the parameter pair (A, *T*_2_) influences the k-space trajectory and affects both scan time and image SNR. As *A* increases, the SPI region decreases and is replaced by radial acquisitions which lead to higher k-space data density and thus a reduction of image noise variance (27). However, the data points experience stronger *T*_2_ weighting which translates into a smaller integral of the MTF (Fig. 1) and hence to a smaller PSF main lobe (Fig. 2), leading to decreased voxel intensity. These two effects partly compensate each other. For small decay coefficients they even largely balance and the improvement of HYFI in SNR efficiency can be well approximated as if arising from scan time reduction alone. High-resolution imaging of short-*T*_2_ components benefits from the use of high gradients (39). As shown in Fig. 6, bone tissue with *T*_2_ ≈ 150 µs can be imaged at an isotropic resolution of 400 µm using a gradient strength of 200 mT/m. Such high gradients induce high bandwidths and thus large k-space gaps, especially in large field of views as required for imaging in humans (Fig. 7). In such circumstances, substituting a large part of the SPI region by radial spokes with HYFI particularly improves scanning efficiency. Moreover, at large gradient amplitude Cartesian SPI acquisition can produce significant mechanical vibrations and acoustic noise due to partly large gradient switching between k-space directions. With HYFI, as long as the gradient can be used at full duty cycle (i.e. without switching back to zero), these effects are significantly reduced since k-space directions are uniformly distributed in all directions and sequentially accessed along a spiral trajectory (40) requiring slower gradient switching, thus clearly improving patient comfort.

The results of this work indicate the potential of HYFI for direct imaging of ultra-short-*T*_2_ components such as in the myelin bilayer in the brain. However as observable in Fig. 7, basic ZTE sequences lead to mostly proton density-weighted images and some kind of selectivity is required to isolate the tissues of interest. One possibility to achieve *T*_2_ selection uses post-processing of a series of images acquired after different dead times. An example of such an experiment is shown in Fig. 9, where *T*_2_ mapping of MnCl_2_ solutions was performed by fitting exponential signal decays. This kind of approach has been shown to enable *T*_2_ selection of ultra-fast relaxing MR signals in the brain which can potentially be assigned to the myelin bilayer (10). Further improvements in quantification with HYFI-based *T*_2_ mapping are expected with a more advanced definition of TE or signal models taking into account the precise sequence timing.

Clinical scanners with state-of-the-art hardware specifications (e.g. RF switching time ∼30 μ, (41), gradient slew rate ∼200 mT/m, and maximum gradient strength ∼80 mT/m, yet at limited duty cycle (17)) may be used for short-*T*_2_ PETRA imaging and will lead to gap sizes of about 30-40 Nyquist dwells. Thus, assuming a TR of a few milliseconds, the acquisition of the inner k-space takes several minutes (c.f. Fig. 2). In such a situation and as illustrated in this paper, substantial improvement in SNR efficiency can be expected when using HYFI instead of PETRA. If the same scanners are used with lower bandwidth (e.g. G < 40 mT/m), the advantage of HYFI is limited to lower acoustic noise and reduced mechanical vibrations. In the special case of a combination of low gradients and large dead times (e.g. G = 10 mT/m, RF switching time ∼50 μ,), the dead time could be used to ramp up the gradient to its target strength, thus allowing smaller excitation bandwidths at reduced gap sizes and avoiding the need of HYFI, yet at the price of reduced spatial resolution (27). A similar idea was exposed in the ramped hybrid encoding (RHE) technique (42), where the readout gradient is lowered during RF excitation and increased to full strength afterwards during data acquisition. In such cases, k-space calibration is required since timing errors and eddy current effects distort the k-space trajectory. Other alternatives to PETRA and hence HYFI are SWIFT (43) and cSWIFT (44), where gaps are very small or inexistent, respectively. However, the first one comes at an SNR penalty and limited bandwidth (45) and the second approach is particularly sensitive to RF coil loading variations.

Finally, there are situations where a large part of the data should be acquired within a small time range as required for chemical shift artifact reduction or *T*_2_ mapping. For example, for performing the latter, longer dead times should be used to include the complete k-space in the gap as performed in Fig. 9, leading to a pure SPI acquisition with spherical support in the PETRA case. Note that in this context, gradients should be switched on *before* the RF pulse since large gaps are actually targeted. Hence, even with clinical scanners, HYFI can be considered as an efficient alternative for SPI methods (24,46).

## 5 Conclusion

The HYFI technique provides both high SNR efficiency and image quality, thus outperforming previously known ZTE-based pulse sequences. It is particularly advantageous in situations involving large dead times or high gradient strengths where PETRA suffers from long and noisy SPI acquisitions. Promising applications include direct imaging of ultra-short *T*_2_ components, such as e.g. the myelin bilayer or collagen, *T*_2_ mapping of ultra-fast relaxing signals, and ZTE imaging with reduced chemical shift artifacts.

## Data Availability

Matlab codes exemplifying the HYFI reconstruction algorithm are available at the cited DOI.

https://doi.org/10.3929/ethz-b-000415045

## Abbreviation

ZTE: Zero echo time
PETRA: Pointwise encoding time reduction with radial acquisition
HYFI: ZTE MRI with hybrid filling
SPI: Single point imaging

## Supporting Information captions

Figure S1: 1D point spread functions (PSFs) for varying decay coefficients A. Simulations were performed for a matrix size of 200 pixels with T_2_ = 100 Nyquist dwells, k_gap_ = 30 Nyquist dwells. For small values of A, the PSFs are very similar. However, the size of the side lobe quickly increases as A approaches 1.

Figure S2: Profiles of the 3D simulations illustrated in Fig. 4 demonstrating the effects of the decay coefficient A on image quality. Each profile is taken horizontally on the middle line of 4 selected images (A = 0, 0.04, 0.1, 0.2). As A increases, the overall image intensity decreases (by a factor smaller than A) and artifacts start to appear in the center of each object, especially in the largest one. Noticeably, the sharpness of the edges is hardly affected. All of these effects can be understood by looking at the HYFI MTF (Fig. 1e). First, as long as the overall PSF shape remains close to a delta function, the image intensity is related to the PSF maximum and hence to the integral of the MTF. As A increases, the MTF decreases by a factor A only in the inner k-space at the end of each radial shell. Thus, its integral and hence the image intensity decreases, but by a factor smaller than A. Second, the irregularities in the MTF arise around the k-space center and not on the edges of the k-space support (as opposed to usual Gibbs ringing). Hence, the related artifacts are expressed as low frequency modulation appearing essentially at the center of the larger objects where PSFs of surrounding pixels can constructively interfere. However, the high-frequency part of the object and hence the resolution of the edges remain mostly unchanged.

